# Nirmatrelvir/ritonavir and molnuipiravir in the treatment of mild/moderate COVID-19: results of a real-life study

**DOI:** 10.1101/2022.08.23.22278585

**Authors:** Ivan Gentile, Riccardo Scotto, Nicola Schiano Moriello, Biagio Pinchera, Riccardo Villari, Emilia Trucillo, Luigi Ametrano, Ludovica Fusco, Giuseppe Castaldo, Antonio Riccardo Buonomo, Federico II COVID team

## Abstract

Molnupiravir and Nirmatrelvir are the first available oral antivirals (OA) active against SARS-CoV-2. However, the trials evaluating the efficacy of OAs involved patients unvaccinated and infected with variants different from those currently circulating. The purpose of this study is to provide real-life data on the efficacy and safety of OAs during the omicron surge of COVID-19 pandemic in a cohort of mostly vaccinated patients.

We conducted a retrospective study on patients with confirmed SARS-CoV-2 infection treated with OAs during the omicron surge in Italy.

We enrolled 257 patients. Of these, 146 (56.8%) were treated with molnupiravir and 111 (43.2%) with nirmatrelvir/ritonavir. Patients in molnupiravir group were older, had a lower body mass index, and a higher rate of chronic heart disease than those treated with nirmatrelvir/ritonavir.

During the 14-day follow-up, four hospitalizations were recorded (1.6%), three in molnupiravir (2.1%) and 1 in nirmatrelvir/ritonavir (0.9%) group. Only one patient (who had received molnupiravir) died. Median time-to-negativity of nasal swab was 8 days (8 days in nirmatrelvir/ritonavir *vs*. 10 days in molnupiravir group, p<0.01).

Globally, we recorded 37 adverse drug reactions (mainly dysgeusia, diarrhea, and nausea) in 31 of 257 individuals (12.1%). Only two patients (0.8%), both receiving molnupiravir, terminated treatment due to the development of adverse drug reactions.

In conclusion, during the omicron surge, in a population of mostly vaccinated patients treated with molnupiravir or nirmatrelvir/ritonavir, we observed a low rate of hospitalization, death, and adverse drug reactions. These rates were even lower than those reported in pivotal trials.

## Introduction

Covid 19 is a disease caused by the betacoronavirus SARS-CoV-2 (1). In late 2019 in Wuhan, China, the first instances of Covid-19 were discovered (2), and the virus quickly spread around the globe(3) causing more than 500 million infections and 6 million fatalities to date (4).

Antivirals against SARS-CoV-2 work by inhibiting viral replication and prevent, in most cases, deterioration of patients towards a severe form of the disease (5).

Several antivirals active against COVID-19 are currently available. The first one to be widely used was remdesivir, a polymerase inhibitor which is administered intravenously. However, this drug has logistical issues because it requires intravenous injections (6). Monoclonal antibodies are another category of drugs which inhibit virus entry into the host cell and are to be administered intravenously as well.

However, the real revolution in the treatment of COVID-19 was the introduction in clinical practice of orally available antivirals Molnupiravir and Nirmatrelvir. (7, 8).

Molnupiravir acts by binding to the RNA-dependent RNA-polymerase causing multiple errors leading to a “lethal mutagenesis” and finally blocking viral replication (9-11).

Nirmatrelvir acts by inhibiting the viral protease Mpro which is essential for viral replication (12, 13). It is administered with ritonavir to boost his pharmacokinetics, a CYP3A4 inhibitor (12).

With respect to registration trials of oral antivirals, it is noteworthy that patients infected with the omicron variant were not enrolled. However, Molnupiravir and Nirmatrelvir should not be affected by mutations that occur mainly in the spike protein, because this protein is not the target of the two drugs. This theoretical consideration is supported by *in vitro* studies which show similar susceptibility to the two antivirals of BA.2.12.1, BA.4, and BA.5 compared to the original Wuhan strain (14). This finding, anyway, needs to be confirmed by *in vivo* data.

Indeed, very few studies on the real-life efficacy of oral antivirals (OA) against omicron variant and in vaccinated individuals are currently available.

The present study aims to provide real-life data on the efficacy and safety of Molnupiravir and Nirmatrelvir/ritonavir during the omicron surge of COVID-19 pandemic.

## Methods

This retrospective real-life study was conducted among all patients referring to the Unit of Infectious Diseases, University of Naples Federico II, Campania Region, Italy, between 18^th^ of February 2022 to 30^th^ of June 2022 with a diagnosis of SARS-CoV-2 infection who were treated with oral antivirals for COVID-19, namely molnupiravir or nirmatrelvir/ritonavir.

Diagnosis of SARS-CoV-2 infection was made through an antigenic or PCR-based validated test.

No exclusion criteria were set, in order to provide real-life results not influenced by selection criteria. However, in Italy the administration of OAs for COVID-19 is regulated by strict indications provided by the Italian Drug Agency (AIFA, *Agenzia Italiana del Farmaco*) (15). In particular, patients with SARS-CoV-2 infection eligible for OA treatment are those not requiring hospitalization due to COVID-19 and at high risk for disease progression due to older age and comorbidities; OAs must be administered within 5 days from COVID-19 symptoms onset. Comorbidities conferring an increased risk of COVID-19 progression include active onco-haematological disease; chronic kidney disease (CKD); severe chronic lung disease; primary or acquired immunodeficiency; obesity (body mass index [BMI]) ≥ 30; severe cardiovascular disease; uncontrolled diabetes mellitus.

All patients referred for OAs were screened for indication by a dedicated medical site staff, which was also responsible for the choice of treatment. All patients were screened for drug-to-drug interactions (DDI) between concomitant chronic treatments and nirmatrelvir/ritonavir. In the presence of significant DDIs, patients were treated with molnupiravir, otherwise nirmatrelvir/ritonavir was the treatment of choice in most cases. OAs were administered for five days at a dosage of 300/100 mg of nirmatrelvir/ritonavir twice daily or 800 mg of molnupiravir twice daily. In case of nirmatrelvir/ritonavir administration, patients with an eGFR between 30 and 60 ml/min/m^2^ received 150/100 of nirmatrelvir/ritonavir twice daily. All treated patients with an eGFR lower than 30 ml/min/m^2^ received molnupiravir.

All eligible patients were asked to sign a specific consent form, as requested by AIFA, and to undergo a medical examination before receiving the assigned treatment (Day 0). All patients received detailed instructions on how to take OAs and were asked to autonomously assume treatment at home. Patients were also trained on identifying adverse drug reactions (ADR) possibly related to OAs and were asked to take note of all symptoms possibly occurring during and after treatment. They were also invited to contact the medical staff in case of necessity and to promptly notify them of the finding of a negative SARS-CoV-2 swab. Aside from spontaneous contact of included patients, the medical site staff performed a telephonic evaluation for all treated patients on Day 7 and Day 14 to investigate the occurrence of new COVID-related symptoms and possible ADRs. A COVID-related hospitalization was defined as the need for hospitalization in patients treated with OAs for SARS-CoV-2 infection, requiring oxygen supplementation treatment for progression of COVID-related symptoms. All causes hospitalizations were recorded as well.

The study’s endpoints were to assess COVID-19 related and all-causes hospitalization rate and mortality rate among patients with SARS-CoV-2 infection treated with OAs according to AIFA criteria. The incidence of ADR was also recorded, as well as risk factors for hospitalization.

### Statistical Analysis

All the variables were tested for parametric/non-parametric distribution with the Kolmogorov-Smirnov test. Comparisons between categorical dichotomous variables were performed with the χ^2^ test (or with Fischer’s exact test when applicable), while comparisons between ordinary variables were conducted with the t-student test (parametric variables) or the Mann-Whitney’s U test (non-parametric variables). Hospitalization rate was reported as person-per-year (PPY) with 95% confidence interval (95CI). All the statistics were performed with IBM^©^ SPSS software, version 26.

## Results

We enrolled 257 patients with SARS-CoV-2 infection who were treated with OAs for COVID-19 during the study period. Among these, 146 (56.8%) were treated with molnupiravir, while 111 (43.2%) received the nirmatrelvir/ritonavir combination.

Clinical data of enrolled patients are shown in Table 1.

**Table 1.**
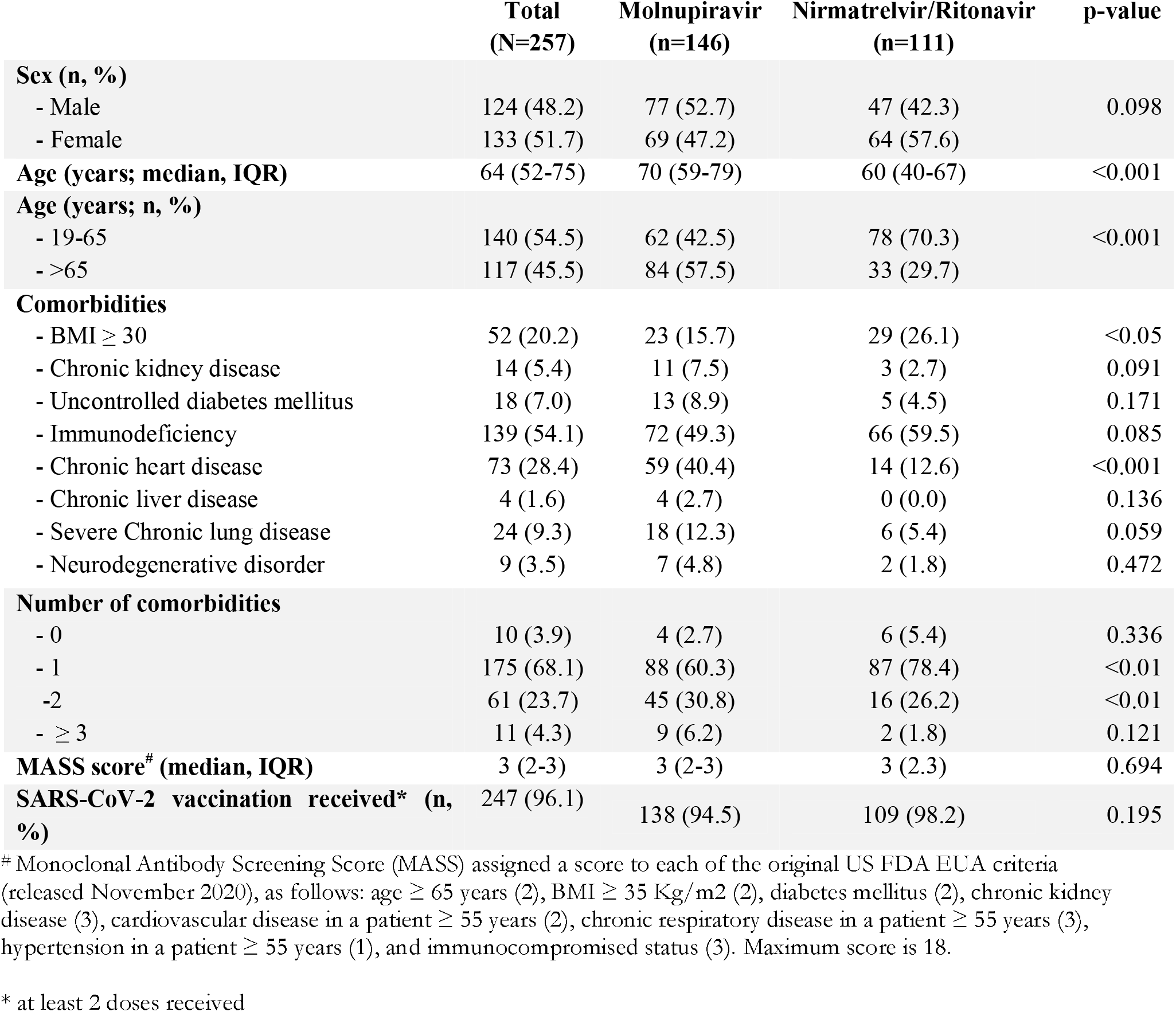
Clinical characteristics of patients with SARS-CoV-2 infection who received oral antiviral therapy

|  | **Total**<br>**(N=257)** | **Molnupiravir**<br>**(n=146)** | **Nirmatrelvir/Ritonavir**<br>**(n=111)** | **p-value** |
| --- | --- | --- | --- | --- |
| **Sex (n, %)** |  |  |  |  |
| - Male | 124 (48.2) | 77 (52.7) | 47 (42.3) | 0.098 |
| - Female | 133 (51.7) | 69 (47.2) | 64 (57.6) |  |
| **Age (years; median, IQR)** | 64 (52-75) | 70 (59-79) | 60 (40-67) | <0.001 |
| **Age (years; n, %)** |  |  |  |  |
| - 19-65 | 140 (54.5) | 62 (42.5) | 78 (70.3) | <0.001 |
| - >65 | 117 (45.5) | 84 (57.5) | 33 (29.7) |  |
| **Comorbidities** |  |  |  |  |
| - BMI $\geq 30$ | 52 (20.2) | 23 (15.7) | 29 (26.1) | <0.05 |
| - Chronic kidney disease | 14 (5.4) | 11 (7.5) | 3 (2.7) | 0.091 |
| - Uncontrolled diabetes mellitus | 18 (7.0) | 13 (8.9) | 5 (4.5) | 0.171 |
| - Immunodeficiency | 139 (54.1) | 72 (49.3) | 66 (59.5) | 0.085 |
| - Chronic heart disease | 73 (28.4) | 59 (40.4) | 14 (12.6) | <0.001 |
| - Chronic liver disease | 4 (1.6) | 4 (2.7) | 0 (0.0) | 0.136 |
| - Severe Chronic lung disease | 24 (9.3) | 18 (12.3) | 6 (5.4) | 0.059 |
| - Neurodegenerative disorder | 9 (3.5) | 7 (4.8) | 2 (1.8) | 0.472 |
| **Number of comorbidities** |  |  |  |  |
| - 0 | 10 (3.9) | 4 (2.7) | 6 (5.4) | 0.336 |
| - 1 | 175 (68.1) | 88 (60.3) | 87 (78.4) | <0.01 |
| - 2 | 61 (23.7) | 45 (30.8) | 16 (26.2) | <0.01 |
| - $\geq 3$ | 11 (4.3) | 9 (6.2) | 2 (1.8) | 0.121 |
| **MASS score<sup>#</sup> (median, IQR)** | 3 (2-3) | 3 (2-3) | 3 (2.3) | 0.694 |
| **SARS-CoV-2 vaccination received\* (n, %)** | 247 (96.1) | 138 (94.5) | 109 (98.2) | 0.195 |

<sup>#</sup> Monoclonal Antibody Screening Score (MASS) assigned a score to each of the original US FDA EUA criteria (released November 2020), as follows: age $\geq 65$ years (2), BMI $\geq 35$ Kg/m<sup>2</sup> (2), diabetes mellitus (2), chronic kidney disease (3), cardiovascular disease in a patient $\geq 55$ years (2), chronic respiratory disease in a patient $\geq 55$ years (3), hypertension in a patient $\geq 55$ years (1), and immunocompromised status (3). Maximum score is 18.
\* at least 2 doses received

Patients treated with nirmatrelvir/ritonavir were younger compared with those treated with molnupiravir (p<0.001) and showed a higher percentage of obesity, among the comorbidities (26.1% vs 15.7%, p<0.05). On the contrary, patients treated with molnupiravir showed a higher percentage of chronic heart disease compared with those treated with nirmatrelvir/ritonavir (40.4% vs 12.6%, p<0.001). Patients with ≥ 2 comorbidities were more frequently treated with molnupiravir than with nirmatrelvir/ritonavir

Throughout the 14-days follow-up, only 4 hospitalizations were recorded among the 257 patients treated with OAs (1.6%). The standardized rate of hospitalizations was 5.68 PPY. Most of the hospitalizations occurred among patients treated with molnupiravir (3, 2.1%; Standardized rate: 7.5 PPY), while only 1 patient (0.9%) treated with nirmatrelvir/ritonavir was hospitalized (Standardized rate: 3.28 PPY). All recorded hospitalization were related to COVID-19 symptoms, thus there was no difference between COVID-related hospitalizations and all-causes hospitalizations. Clinical characteristics of the four patients who needed hospitalization are sown in *Table 2*. Three patients (2.1%) treated with Molnupiravir, and one patient treated with Nirmatrelvir/ritonavir (0.9%) needed hospitalization (p=0.460). Similar rates of hospitalization were recorded in patients with ≥ 2 comorbidities (1.4%) compared with patients with none or 1 comorbidity (1.6%, p=1.000).

**Table 2.** Clinical characteristics of patients who needed hospitalization

| **Patient n°** | **Age**<br>**range**<br>**(years)** | **Sex** | **Comorbidities** | **MAS**<br>**S**<br>**score** | **SARS-CoV-2**<br>**vaccination** | **Treatment received** | **Outcome** |
| --- | --- | --- | --- | --- | --- | --- | --- |
| **1** | 81-85 | M | Immunodeficiency; chronic heart disease; | 3 | Yes | Molnupiravir | Exitus |
| **2** | 76-80 | F | Chronic heart disease | 2 | Yes | Molnupiravir | Recovery |
| **3** | 86-90 | M | Chronic heart disease | 2 | Yes | Molnupiravir | Recovery |
| **4** | 40-45 | M | Immunodeficiency | 3 | Yes | Nirmatrelvir/Ritonavi | Recovery |

Only one patient treated with molnupiravir died. This patient was a men in his 80s with immunodeficiency related to a chronic lymphocytic leukaemia and chronic heart disease due to prior acute coronary syndrome. The median time to obtain a negative SARS-CoV-2 swab in patients treated with OAs was of 8 days (IQR: 7-13), with a lower time in patients treated with nirmatrelvir/ritonavir (8 days; IQR: 6-11) compared with patients treated with molnupiravir (10 days; IQR: 7-17, p<0.01).

With respect to ADRs, globally, we recorded 37 ADRs occurring in 31 of the 257 patients (12.1%), with 26 patients (10.1%) referring one ADR and 5 patients (1.9%) experiencing 2 ADRs. Thirteen patients (8.9%) and 18 patients (16.2%) treated with molnupiravir and nirmatrelvir/ritonavir, respectively, reported at least one ADR (p=0.075). The most common ADR was dysgeusia, reported by 14 patients (5.4%). Other recorded ADRs were: nausea (6 patients, 2.3%); diarrhoea (6 patients, 2.3%); headache (4 patients, 1.6%); skin rash (2 patients, 0.8%); vomit (1 patient, 0.4%); dizziness (1 patient; 0.4%); seizure (1 patient, 0.4%). Only two patients (0.8%), both treated with molnupiravir, discontinued the treatment due to the occurrence of ADRs (seizures and dizziness, respectively). Dysgeusia was more commonly reported by patients treated with nirmatrelvir compared with those who received molnupiravir (9.0% vs 2.7%, p<0.05; *Table 3*).

**Table 3.** Side effects recorded among the included patients and according to treatment received

|  | **Molnupiravir**<br>**(n=146)** | **Nirmatrelvir/Ritonavir**<br>**(n=111)** | **p-value** |
| --- | --- | --- | --- |
| **Diarrhea** | 4 (2.7%) | 2 (1.8%) | 0.701 |
| **Dizziness** | 1 (0.7%) | 0 (0.0%) | 1.000 |
| **Dysgeusia** | 4 (2.7%) | 10 (9.0%) | **<0.05** |
| **Headache** | 2 (1.7%) | 2 (1.8%) | 1.000 |
| **Nausea** | 2 (1.7%) | 4 (3.6%) | 0.407 |
| **Seizure** | 1 (0.7%) | 0 (0.0%) | 1.000 |
| **Skin rash** | 1 (0.7%) | 1 (0.9%) | 1.000 |
| **Vomit** | 1 (0.7%) | 0 (0.0%) | 1.000 |
| **Discontinuation of treatment** | 2 (1.7%) | 0 (0.0%) | 0.507 |

## Discussion

In this prospective, real-life cohort study, 257 patients with SARS-CoV-2 infection were treated with oral molnupiravir or oral nirmatrelvir/ritonavir according to the indications provided by AIFA (15). The introduction of OAs was a revolution in the treatment of COVID-19. However, the pivotal trials which led to their approval enrolled patients that are different from those who could benefit from these drugs mainly with respect to vaccination status and viral variant.

What do we know from clinical trials? Molnupiravir efficacy and safety were assessed in the MOVe-OUT a phase 2/3 randomized, double blind, placebo-controlled trial. This trial enrolled patients with a SARS-CoV-2 infection with symptoms onset within 5 days, and at least one risk factor for development of severe illness from Covid-19: age >60 years; active cancer; chronic kidney disease; chronic obstructive pulmonary disease; obesity, serious heart conditions. The most prevalent variant was delta, followed by mu and gamma. Notably, no omicron cases were enrolled (the trial ended before the first case of the omicron variant) (16). All participants in the trial were not vaccinated. Molnupiravir met the superiority criterion *vs*. placebo: the patients receiving Molnupiravir had a lower risk of death or hospitalization at day 29 after administration compared to those receiving placebo (6.8% vs. 9.7%; difference,−3.0 percentage points; 95% CI, −5.9 to −0.1) (16). Another trial has been conducted to assess the utility of Molnupiravir in patients requiring in-hospital treatment for Covid-19 with symptom onset ten or fewer days before randomization. In this context, the drug failed to manifest any clinical benefit (17).

The efficacy of the combination of Nirmatrelvir and Ritonavir was evaluated in the EPIC-HR trial: a double-blind, randomized, placebo-controlled trial(18). This trial enrolled adults older than 18 years with at least one risk factor for progression to severe disease with a confirmed SARS-CoV-2 infection and symptom onset not older than 5 days. All patients in the trial were not vaccinated and did not experience an earlier SARS-CoV-2 infection. The authors of the trial did not report a detail of SARS-CoV-2 variants. However, the trial enrolment finished just a few days after the first omicron cases were diagnosed in the world (19), so omicron prevalence among the trial subjects should be minimal, if any. Primary objective of the trial was to assess the drug’s efficacy in preventing Covid-19 related hospitalization or death. The drug successfully reached the endpoint, resulting superior to placebo (difference of −5.81 percentage points 95% CI, −7.78 to −3.84; P<0.001; relative risk reduction, 88.9%) (18).

In consideration of the study period (February-June 2022), included patients were assumed to mostly harbour the omicron variant of SARS-CoV-2. Given the eligibility criteria for OA treatment, most patients included in this analysis were aged or had significant comorbidities. This evidence represents a substantial difference with clinical trials investigating efficacy and safety of OAs for SARS-CoV-2. In fact, the median age of enrolled patients in the pivotal trial of nirmatrelvir/ritonavir and molnupiravir was 45 (95CI: 18-86) and 42 (18-90) years, respectively (18). Furthermore, in both these clinical trials, increased Body Mass Index was the most common risk factor for COVID-19 progression among the enrolled patients. Obesity was indeed the most frequent comorbidity of patients treated with molnupiravir in the MOVe-OUT trial, while 80.5% of patients enrolled in the EPIC-HR study showed a BMI of 25 or above. On the contrary, a significant percentage of patients included in our real-life cohort had primary or iatrogenic immunodeficiency mostly related to active haematological disease and immunosuppressant treatment (49.3% in the molnupiravir group, 59.5% in the nirmatrelvir/ritonavir group). Only a paucity of patients in the EPIC-HR and in the MOVe-OUT trials had immunodeficiency, overall making the population of the two trials considerably different from the real-life population of our study. Results from other real-life cohorts and retrospective studies assessing safety and efficacy of OAs for SARS-CoV-2 also refer to populations with a very low rate of patients with immunodeficiency, with obesity generally representing the most common risk factor for COVID-19 progression (20, 21). These differences must be taken into account when efficacy and safety data of OAs from this cohort are reported.

Despite the high rate of patients with advanced age and severe comorbidities, the outcome was favourable in most cases. Only 4 patients (1.6%, 5.68 PPY) indeed needed hospitalization and only 1 patient (0.4%) died, without significant differences between the two antivirals. Even without an untreated control group, it is noteworthy that the rates of hospitalization and death were considerably lower compared with those reported in clinical studies and by governmental agencies. Results from the COVID-NET network indeed showed that, in a period of omicron predominance, the proportion of hospitalized, vaccinated adults with COVID-19 peaked at 13.4% among the general population (22). We underline that our cohort includes a high number of patients with immunodeficiency who are, among at-risk patients, those particularly at risk for severe COVID-19, hospitalization and death (23-26).

Also comparing our data with those that emerged from pivotal trials, it is noteworthy that the rate of hospitalization reported among patients treated with molnupiravir in our cohort was consistently lower compared with the hospitalization rate reported in the MOVe-OUT trial (2.1% vs 7.3%) (16), while the rate of hospitalized patients treated with nirmatrelvir/ritonavir was similar to the one recorded in the EPIC-HR study (0.9% vs 0.72%) (18). However, these results should be interpreted in the light of the characteristics of the population of our study, which is made up of nearly all vaccinated, tough frail subjects

Very few real-life studies are available so far. A large study has been conducted in Israel on a healthcare provider’s database on patients e diagnosed with SARS-CoV-2 infection between January and February 2022, who were at high risk for severe COVID-19 and had no contraindications for nirmatrelvir/ritonavir use (20). On the total sample of over 180,000 eligible patients. 4,732 (2.6%) were treated with nirmatrelvir/ritonavir. The authors found a significant decrease in the rate of severe COVID-19 (adjusted HR 0.54; 95% CI, 0.39-0.75) or death (adjusted HR 0.20; 95% CI, 0.17-0.22) for the patients treated with nirmatrelvir/ritonavir with respect to untreated patients(20). The majority of patients (75.1%) were vaccinated against SARS-CoV-2. Information about the variants was not available, but according to the timeframe in which the study was performed, it is likely that at least in the second half of the study BA.2 was the predominant variant (20). Another retrospective study was conducted in Hong Kong. This study evaluated all-cause mortality in hospitalized patients with a mild form of COVID-19 receiving molnupiravir or nirmatrelvir/ritonavir. Each group was matched in a ratio of 1:1 with untreated controls. The study found that the use of both OAs was associated with a reduction in all cause mortality: molnupiravir HR=0.48 (HR=0.48, 95%CI=0.40-0.59, p<0.0001); nirmatrelvir/ritonavir: HR=0.34 (95%CI=0.23-0.50, p<0.0001) (27). Mortality rates were 8.1% and 15.9% in the molnupiravir and control group, respectively and 3.6% and 10.3% in the nirmatrelvir/ritonavir and control group, respectively. However, differently from our cohort, only a minority of the patients in that study were vaccinated: 6.2% of those receiving molnupiravir and 10.5% of those receiving nirmatrelvir/ritonavir.

Another interesting result of our study was the tolerability of oral antivirals. Despite the old age of included patients and the presence of severe comorbidities, the rate of recorded ADRs was low. In fact, only 12.1% of patients treated with OAs reported an ADR (8.9% in the molnupiravir group and 16.2% in the nirmatrelvir/ritonavir group). Furthermore, the rates of patients with ADR were lower compared with data of the MOVe-OUT trial (30.4% of patients with at least one ADR in the molnupiravir group) (16) and of the EPIC-HR trial (22.6% of patients with at least one ADR in the nirmatrelvir/ritonavir group) (18). Interestingly, the most common ADR reported from patients in this cohort was dysgeusia. This ADR was more frequent among patients treated with nirmatrelvir/ritonavir compared with those treated with molnupiravir (9.0% vs 2.7%, p<0.05), while the occurrence of dysgeusia was not reported in the MOVe-OUT trial (16). Otherwise, in the EPIC-HR trial, 5.3% of patients treated with nirmatrelvir/ritonavir experienced dysgeusia (18), confirming a possible association between dysgeusia and treatment with nirmatrelvir/ritonavir. We acknowledge that clinical trials usually assess ADRs differently and often more thoroughly compared to real-life studies. However, we underline the very low rate of discontinuation of our cohort which reflects an excellent tolerability of both antivirals.

The strength of this work is the availability of real-life data in vaccinated patients during the omicron surge and the assessment of safety in a cohort of patients aged and affected by severe comorbidities, in particular immunodeficiency. On the contrary, a consistent limit of this study is represented by the absence of a control group of untreated patients. However, it must be said that denying OA treatment to frail patients with SARS-CoV-2 in a real-life scenario should be considered unethical. Another limit of our study is represented by the absence of a systematic follow-up at the medical site to assess viral clearance. Data on follow-up swabs for SARS-CoV-2 were mostly spontaneously reported by the included patients.

In conclusion, results from this real-life retrospective study showed a very low rate of hospitalization, death and ADRs among patients with SARS-CoV-2 treated with oral molnupiravir or nirmatrelvir/ritonavir. Rates of such outcomes were lower compared with those reported in randomized controlled trials and in other real-life experiences with OAs for SARS-CoV-2 infection.

## Data Availability

All data produced in the present study are available upon reasonable request to the authors

